# Design and Evaluation of a Bone-anchored, Neurally-controlled Knee Prosthesis

**DOI:** 10.64898/2026.01.14.26343865

**Authors:** John A. McCullough, Daniel Levine, Tony Shu, Rickard Brånemark, Matthew J. Carty, Hugh M. Herr

## Abstract

**Background:** Commercially-available microprocessor-controlled prosthetic knees are unable to fully replicate the biomechanical function of the missing biological limb. While powered prostheses have the capacity to restore joint level kinetics, current systems rely on intrinsic control schemes that do not allow the user to volitionally modulate movement under neural commands. This limitation may compromise functional performance and hinder prosthetic embodiment, the sense that the device is part of the user’s body. In a case study on one test participant, we evaluate the functional and perceptual benefits of a bone-anchored, neurally-controlled knee prosthesis by comparing it to the participant’s microprocessor-controlled prosthesis.

**Methods:** We conducted a within-subject study on an individual with a transfemoral amputation, with an osseointegrated implant and surgically reconstructed agonist–antagonist muscle pairs. We tested a neurally-controlled powered knee and conventional microprocessor knee across a set of activities, including seated volitional control tasks, sit-to-stand transitions, squatting, level-ground walking, stair ascent, and uninstructed standing. Performance metrics included knee kinematics, prosthesis-generated mechanical power, and functional outcomes such as gait speed, stair ascent time, and weight-bearing symmetry derived from ground reaction forces. Functional mobility and control were complemented by self-reported embodiment, assessed through a questionnaire targeting agency, ownership, and body representation.

**Results:** The neurally-controlled prosthesis enabled intuitive and responsive control. Compared to the subject’s prescribed prosthesis, the prosthesis yielded improved temporal gait symmetry during walking (symmetry index: 0.93 vs. 0.59, with 1 indicating perfect stance time symmetry), increased prosthetic-side weight-bearing during sit-to-stand and squatting, and successful execution of a step-over-step stair ascent strategy—an outcome not achievable with the subject’s prescribed device. Embodiment scores were consistently higher with the neurally-controlled prosthesis compared to the prescribed device across multiple domains, including agency, ownership and body representation.

**Conclusions:** This study is the first to directly compare a prescribed microprocessor knee with a bone-anchored, neurally-controlled powered prosthesis. By combining osseointegration, surgically reconstructed agonist–antagonist muscle pairs, and powered actuation, the system improved gait symmetry, greater prosthetic-side loading, and step-over-step stair ascent. These results demonstrate the novelty and promise of integrating surgical and mechatronic innovations to restore both functional mobility and embodied control after transfemoral amputation.

**Trial registration:** This study was approved by the Institutional Review Board at MIT (Protocol No. 2503001589).

## Background

Transfemoral amputation results in the loss of multiple joints critical for coordinated, economical movement, making tasks such as sit-to-stand transitions, stair ascent, and uneven terrain navigation significantly more challenging [1], [2], [3]. Most individuals with transfemoral amputation rely on either passive or quasi-passive prosthetic knees, the latter typically comprising microprocessor-controlled variable dampers. These variable-damper knees adjust resistance based on gait phase to enhance stability and safety but cannot generate positive mechanical power [4]. As a result, this class of device cannot replicate the full biomechanical function of the biological knee, often leading to compensatory strategies such as increased reliance on the intact limb and asymmetrical gait patterns [3], [5], [6]. Tasks that require active knee extension, such as stair ascent, remain particularly difficult, as these quasi-passive devices cannot generate the necessary positive mechanical power [7].

Powered prostheses address these limitations by incorporating electromechanical actuators, such as electric motors, to actively generate positive power during ambulation [8], [9]. However, designing effective control systems for powered devices remains a significant challenge. Prosthesis controllers must assist natural human movement while adapting to changing environments in real time. In other words, such powered systems must determine both when and how much power to deliver or regenerate. Most existing prosthetic designs have no integration with the user’s biological control mechanisms, such as the Ottobock Empower Ankle-foot Prosthesis and the Össur Power Knee [10], [11], [12], [13], [14], [15]. Other prosthetic controllers use noninvasive approaches, such as surface electromyography, to detect human intent [16], [17], [18], [19], [20], [21], [22]. Unfortunately, these systems face challenges such as signal drift, impulse artifacts, and cross-talk [23].

In addition to biomechanical limitations, all commercially-available lower-extremity prosthetic devices lack meaningful integration with the user’s nervous system. The lack of communication between the biological systems and hardware results in degraded prosthesis embodiment, defined as the user’s sense of ownership, agency, and body representation with the device [24], [25], [26], [27], [28]. The degree to which a prosthesis is embodied is correlated with device acceptance and continued use. Consequently, prostheses that do not evoke a sense of ownership or agency are more likely to be abandoned, even if they restore some physical function [25], [29], [30]. As such, promoting embodiment is not only a psychological goal, but a practical one with significant implications for long-term outcomes.

Recent advances in surgical reconstruction techniques and myoneural interfacing strategies offer promising new opportunities for direct human-prosthesis integration [31]. The Agonist-antagonist Myoneural Interface (AMI) is a surgically-constructed muscle pairing that restores natural proprioceptive feedback and enables efferent motor intent sensing to control external devices [32], [33]. Previous work has demonstrated that the AMI using surface electromyography (sEMG) can support continuous neural control of a powered ankle and restore biomimetic gait patterns [34], [35]. When AMIs are coupled with osseointegrated implants enabling intramuscular electrodes (e.g., the eOPRA Implant System, Integrum AB, Mölndal, Sweden), users with transfemoral amputation have demonstrated volitional control over a powered knee prosthesis [36]. In this preliminary case study on one test participant, we extend the work of Shu *et al*. [36] to evaluate the functional and perceptual benefits of a bone-anchored, neurally-controlled knee prosthesis. We hypothesize that volitional control with AMI muscle signals from implanted electrodes will improve biomimetic performance and enhance the user’s sense of embodiment compared to when a commercially-available, microprocessor knee is employed. We tested the biomimetic performance aspect of this hypothesis by conducting experiments as the subject used both the neurally-controlled prosthesis and their prescribed microprocessor knee prosthesis during free-space movements, sit-to-stand and squatting maneuvers, level-ground walking, stair ascent, and periods of uninstructed standing. Further, the embodiment aspect of the hypothesis was assessed through a 50-item questionnaire.

## Methods

### Prosthesis Design and Control

The powered knee prosthesis used in this study was a modular robotic limb designed to support continuous volitional control via implanted EMG electrodes (Fig. 1). The actuator was based on the design by Carney *et al.* [37], comprising of a brushless DC motor (T-Motor U10 Plus, T-Motor, Nanchang, China) that drove a 5 mm lead ball-screw, coupled to a series-elastic element arranged within a parallelogram linkage. The prosthesis has a nominal gear ratio of 52:1 ± 3.5 and a range of motion (ROM) of approximately 90° [37]. This configuration allowed for the generation of biologically-relevant joint torques and speeds necessary for daily activities of a 100 kg user. The open-loop torque bandwidth of the actuator was 4.5 Hz [36], [37]. The fully integrated device weight was 2.6 kg (Table S1), falling within anthropometric norms for commercially-available prosthetic knees.

**Figure 1:**
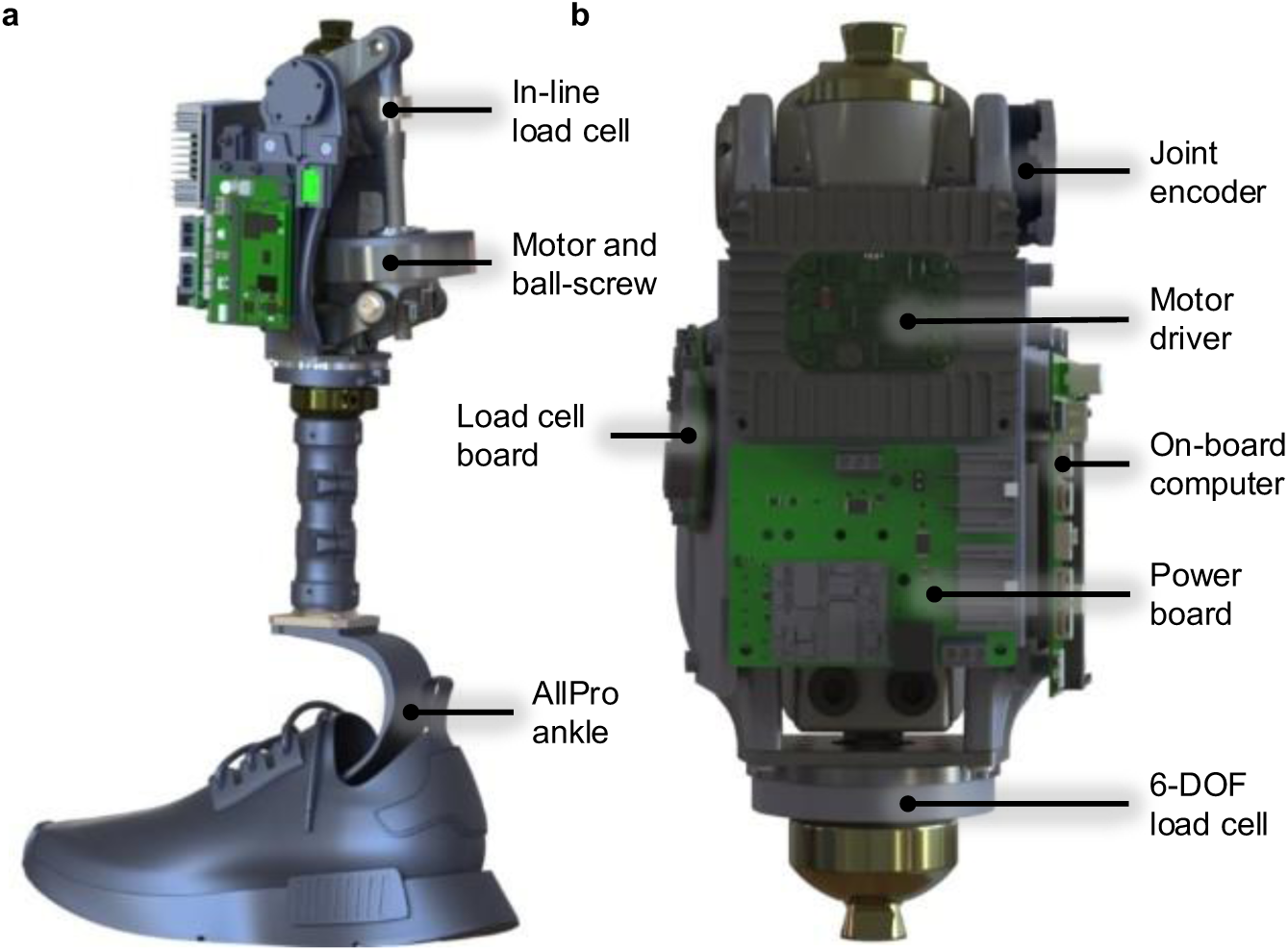
Mechanical and embedded system architecture of the neural-controlled knee prosthesis. **a** Lateral view showing the full prosthesis assembly. **b** Frontal cross-section view highlighting the embedded components.

The actuator was powered by an Ingenia Everest XCR motor driver (Novanta Inc., Beford, MA), which received current commands at 1 kHz. Safety features such as thermal monitoring, overcurrent protection, and safe torque off were integrated directly into the drive electronics. The motor driver was thermally managed using a custom-designed heat sink for continuous operation (Fig. S7).

An embedded computer (Verdin iMX8MP, Toradex, Lucerne, Switzerland) ran a modular, multithreaded software architecture on an embedded Linux platform (Fig. S2). This computer interfaced with sensors and actuators via a custom carrier board that incorporated two high-resolution analog-to-digital converters (ADCs), an inertial measurement unit (IMU), and a controller area network (CAN) transceiver (Fig. S1). Sensor polling, control execution, and data logging were performed at 1 kHz in real time. CAN object exchange protocol (CANOpen) communication with the motor driver was handled via the Lely stack (Lely Industries, Maassluis, Netherlands), providing low-latency and fault-tolerant performance (Fig. S2). Power was supplied by two 6S1P lithium polymer (LiPo) batteries, which delivered 48 V to the actuator and 12 V to the control electronics via an isolated direct-current to direct-current (DC-DC) converter (Fig. S6). A custom power distribution board routed power to the appropriate subsystems and supports future expansion for additional actuators, battery health monitoring, and user diagnostics (Fig. S6). Peripheral sensors included a high-resolution load cell (Futek LCM300) mounted on the ball-screw to measure motor-side force output, a 6-degree-of-freedom (DOF) load cell (Sunrise Instruments M3564F1) for measuring ground reaction forces, and an absolute magnetic encoder (RLS RM08) for joint angle tracking (Fig. S1). Additional information is provided in Additional file 1.

Electromyographic (EMG) signals were recorded from bipolar intramuscular electrodes implanted in AMI muscle pairs, the rectus femoris (extensor) and biceps femoris (flexor). The electrodes were routed percutaneously via the eOPRA abutment and connected to a custom EMG acquisition system [38]. EMG signals were sampled at 2 kHz and band-pass filtered using a finite impulse response filter (passband: 70–350 Hz; stopbands: 0–60 Hz and >350 Hz; 85 dB attenuation; order 198) to reduce motion artifacts and high-frequency noise [23]. The filtered signals were full-wave rectified and smoothed using a 100 ms root-mean-square moving window. Processed EMG was then normalized, producing a scalar activation signal ranging from 0 to 1. These activation signals were used to generate torque commands for the knee joint.

The neural prosthesis employed a hybrid control architecture that combined intrinsic’ task-invariant behavior with continuous user-driven modulation. A finite state machine (FSM) segmented gait into two phases (stance vs. swing) based on vertical ground reaction forces measured by the 6-DOF load cell (Fig. S3). An intrinsic controller provided baseline impedance-based control, while a neural controller, developed by Shu *et al.* [36], modulated joint torque in real time using processed EMG (Fig. S4). Muscle signals were mapped to joint stiffness, damping, and equilibrium position via logistic and affine transformations, enabling physiologically meaningful impedance modulation. In contrast to the original Shu *et al.* implementation, the intrinsic controller was simplified, and gravitational compensation torque was omitted. Torque commands from both controllers were summed and passed to a closed-loop torque control system (Fig. S5). This system used a feedforward-feedback structure to ensure accurate and stable joint actuation and achieved a closed-loop torque bandwidth of 10 Hz, sufficient for dynamic locomotor tasks [39]. The commanded knee torques were converted to motor currents using a mechanical model that incorporated the linkage moment arm, drivetrain gear ratio, and motor torque constant. For more details on the mechatronic implementation see Additional file 1.

### Experimental Protocol

To evaluate the performance of the neurally-controlled prosthesis, we conducted a comprehensive set of experiments in a single male participant in his 60s with a unilateral transfemoral amputation. The participant was three years post-revision surgery involving the creation of an AMI muscle pair and implantation of an eOPRA osseointegrated endoprosthesis. These surgical constructs enabled bidirectional neuromuscular interfacing via implanted bipolar intramuscular electrodes. The participant had prior experience using neurally-controlled lower-limb prostheses.

A within-subject, crossover design was used to compare the participants’ performance while using their microprocessor prosthesis (Ottobock C-Leg) and the neural knee developed in this study. A battery of tasks was administered to evaluate volitional control, functional performance during daily activities, and perceived embodiment of the device. All data were collected at the MIT Media Lab in Cambridge, Massachusetts. All procedures were approved by the Institutional Review Board number 2503001589, and written informed consent was obtained prior to participation.

Data were collected across six supervised in-laboratory sessions of approximately 4 hours each, conducted on separate days. At the start of the day, the neurally-controlled prosthesis was mounted to the participant’s eOPRA abutment and aligned in the laboratory by a member of the research team with experience in lower-limb prosthetic alignment. Sessions were structured to limit extended training and thereby reduce confounding adaptation effects. For each device and task, except the free-space task, the participant first completed a 5-minute acclimatization period. This period was used to verify appropriate prosthesis functionality for the task and to ensure that the task instructions were clearly understood, after which formal data collection commenced. The neurally-controlled prosthesis was powered only during periods of active experimentation and was switched off between task blocks and during rest breaks provided for participant comfort. Use of the neurally-controlled prosthesis was restricted to these supervised sessions. Outside the laboratory, the participant continued to ambulate with his prescribed microprocessor-controlled prosthesis.

### Free-space Reaching Task

To assess volitional control, we conducted a free-space experiment in which the participant used muscles in his residual limb to control the neurally-controlled prosthesis while seated. Using flexor (rectus femoris) and extensor (biceps femoris) muscles in the residuum, the participant was instructed to align the prosthetic foot with three predefined angular targets under two feedback conditions: blindfolded and sighted. To minimize the effects of training, the blindfolded condition was conducted first and not permitted to use or practice with the neurally-controlled prosthesis prior to testing with the device. In the sighted condition, the blindfold was removed, but all other procedures remained the same. Between conditions, a rest period was provided to minimize the impact of muscle fatigue. The prosthesis was mounted on a stand adjacent to the user and mechanically isolated from them to minimize the ability to feel motor or structural vibrations. Additionally, the participant wore noise-cancelling headphones to minimize auditory cues of the prosthesis.

A rigid beam with a single target marker was placed parallel to the prosthetic leg. Reflective markers were placed on the toe, midsole, and heel of the prosthetic foot. The low, mid, and high targets required aligning the toe, midsole, and heel markers with the beam marker, respectively. These three targets thus represented distinct low-, mid-, and high-angle knee configurations. In order to quantify angular error between the device and the target market, the participant and researcher first manually set the three target angles. While backdriving the prosthesis, the participant stopped the knee joint when the toe, midsole, and heel markers were aligned with the bean marker. These angles were used as the target angle for the low, mid, and high heights.

Within each condition, the participant completed 24 reaching movements, 8 repetitions at each height, with target heights pseudorandomized. Each movement consisted of a sequence of three seconds of movement, three seconds of holding the target position, and three seconds of rest; instruction were provided by an audio track heard through the headphones. Performance was quantified as the average absolute angular error between the prosthetic knee and the target angle during the middle second of the hold phase. EMG activity of both the flexor and extensor muscles was collected, and the extensor EMG was analyzed, as it was the muscle that could activate the knee extension.

### Sit-to-Stand Transitions and Squatting

Functional performance with the microprocessor and neurally-controlled prostheses was assessed using sit-to-stand and stand-to-sit transitions. For sit-to-stand and stand-to-sit transitions, the participant rose from and returned to a standard-height chair while vertical ground reaction forces (GRFs) were recorded from an instrumented treadmill (Bertec Corporation, Columbus, OH). GRF have been used previously to characterize limb loading and reliance [36], [40], [41]. The participant was instructed to use the handrails as needed for safety. For squatting, the participant descended from an upright standing posture to approximately 90° knee flexion, and then returned to standing.

For each device, the participant first completed a 5-minute acclimatization period to ensure appropriate prosthesis functionality and understanding of the task. Sit-to-stand trials in the acclimatization period were collected during the initial exposure to the neurally-controlled prosthesis, following basic safety instructions before formal data collection was started. Short rest periods were provided between trials to minimize the influence of muscle fatigue on performance.

GRFs were used to determine limb loading symmetry throughout the movement cycle. The slope of the linear regression between normalized prosthesis and intact leg support was calculated to quantify support symmetry; a slope of one represented perfect symmetry between legs. Actuator torque and power were computed from the axial force measured by the load cell placed in series with the ballscrew, combined with the known transmission geometry of the knee mechanism. Power was calculated as the product of the actuator torque and measured knee velocity. Joint kinematics were recorded using a joint encoder on the neurally-controlled device, and for other joints via goniometers (Biometrics Ltd., Newport, UK).

### Level-ground Walking

Level-ground walking was assessed on a split-belt instrumented treadmill. With each prosthesis, the participant’s preferred walking speed was identified by starting at a comfortable slow speed and increasing the belt speed in increments of 0.1 m/s. After each increment, the participant was asked whether the speed felt slower, faster, or closest to their natural daily walking speed. The speed at which the participant reported that the treadmill felt most like their normal walking was designated as the preferred walking speed. To confirm this selection, speeds 0.1 and 0.2 m/s above the chosen value were also tested, but the participant reported them as quicker than their typical pace. GRFs were used to segment gait cycles and identify stance phases, enabling computation of a temporal symmetry index based on single-limb support durations [42]. A symmetry index of 1 indicated equal stance duration on both limbs, defined as:

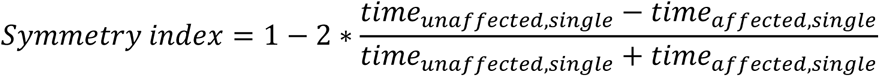

To complement treadmill-based measures, the participant also completed overground walking trials along a 4.88 m walkway at a self-selected pace to assess natural walking speed in the absence of treadmill constraints. Walking speed was computed from the time required to traverse the walkway.

### Stair Ascent

Stair ascent performance was evaluated on a flight of four standard-height, 20 cm, steps. The participant completed five ascent trials at a self-selected pace under each prosthesis condition and was permitted to use handrails for safety. Ascent time was calculated from initial lifting of the prosthetic foot on the first step to final foot placement on the top step. Primary outcome measures included peak prosthetic knee flexion angle during ascent and total ascent duration. In addition to the in-laboratory stair ascent task, the participant also climbed 26 steps outside the laboratory on three occasions.

### Uninstructed Standing

To evaluate uninstructed behavior, we analyzed standing intervals that occurred immediately before the start of walking and squatting tasks. During these brief pre-task periods, the participant was not given any explicit instructions regarding posture or weight distribution. Vertical GRFs were used to compute the regression slope between prosthetic-side and intact-side support over each interval, providing a quantitative measure of how body weight was distributed between limbs during these unprompted standing periods.

### Embodiment Questionnaire

Finally, prosthesis embodiment was assessed using a patient-reported outcome measure (PROM) developed specifically for this study (Table S2). The questionnaire was self-administered by the participant in a private setting without researchers present during completion. The questionnaire consisted of 50 items designed to probe agency, ownership, body representation, phantom sensation, and proprioception in relation to the prosthesis. Agency is the sense of control over one’s movements, ownership is the feeling that the leg or device belongs to one’s body, and body representation is the mental image of one’s body [26]. Phantom sensation was defined as positive, non-painful sensations of the missing limb during ambulation, including awareness of the limb’s presence and position, rather than episodes of phantom pain. Proprioception was defined as the perceived position and movement of the prosthesis as if it were the missing biological limb, independent of visual feedback. Responses were rated on a 5-point Likert scale from 1 (hardly ever) to 5 (almost always), and domain-specific scores were averaged for each device condition. The full questionnaire is provided in Additional file 1. This instrument is a study-specific PROM that has not undergone formal psychometric validation, as such it should be interpreted as an exploratory measure of embodiment within this single-participant case study.

### Statistical Analysis

All statistical analyses were performed using MATLAB R2024b (MathWorks Inc., Natick, MA). For continuous outcomes, we summarized performance using means and standard deviations across repetitions or trials. Embodiment questionnaire responses were summarized as mean scores within each domain for each device. Given that data were obtained from a single participant with repeated measures, no formal hypothesis testing was performed. Instead, observed differences between conditions are interpreted descriptively to characterize the magnitude and consistency of device-related effects.

## Results

### Free-space Volitional Control

The participant demonstrated successful volitional control of the powered knee prosthesis using residual muscle activation across a range of knee extension targets under both blindfolded (Fig. 2a) and sighted (Fig. 2b) conditions. In both conditions, rectus femoris activation scaled appropriately with target height (Fig. 2c). EMG activity was elevated in the blindfolded condition compared to the sighted condition at all target levels, indicating greater muscle effort when visual feedback was unavailable. Task accuracy, quantified as the average absolute angular error between the actual and target knee angles, improved with the addition of visual feedback (Fig. 2d). The errors were lower in the sighted condition for mid and high targets, with a similar trend at the low target. These findings confirm that the participant could generate reliable joint-level control through neural input alone, and that visual feedback improved accuracy and reduced muscular effort.

**Figure 2:**
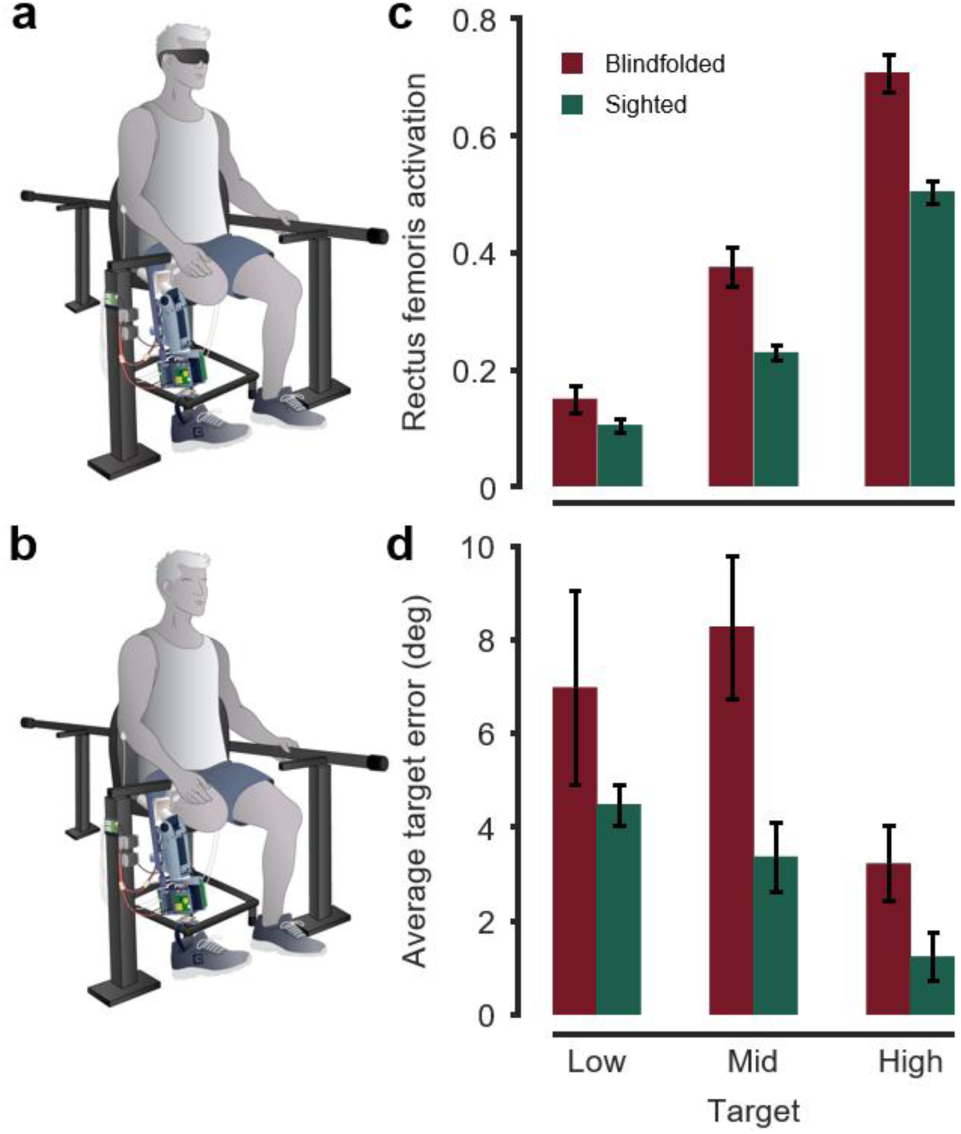
Volitional control of a neural prosthesis using residual muscle activation. Experimental setup for the free-space in **a** blindfolded and **b** sighted conditions. **c** Rectus femoris activation increased with target height and was higher in the blindfolded condition compared to the sighted condition at all target levels, indicating greater effort without visual cues. **d** Average target error decreased in the sighted condition, with lower angular error for mid and high targets. Error bars represent standard error.

### Sit-to-Stand Transitions

The participant exhibited controlled, volitional performance during sit-to-stand and stand-to-sit transitions (Fig. 3a) with both devices. Knee kinematics were smooth and comparable across prescribed and neural conditions (Fig. 3b). Joint torque and power analyses confirmed that the neurally-controlled prosthesis contributed actively during movement, producing positive and negative power during the stand and sit phases, respectively (Fig. 3c). Notably, the peak power at the knee joint was 0.54 W/kg, whereas the biological peak power of young adults is 0.14 W/kg [43]. During the acclimatization period, both flexor and extensor muscles were recruited to coordinate movement, but then the participant transitioned to primarily using rectus femoris activation while suppressing biceps femoris activity (Fig. 3d). Load-sharing analyses indicated greater engagement of the prosthetic limb during the standing phase with the neural prosthesis (Fig. 3e). However, during seated phases, load distribution remained comparable across devices.

**Figure 3:**
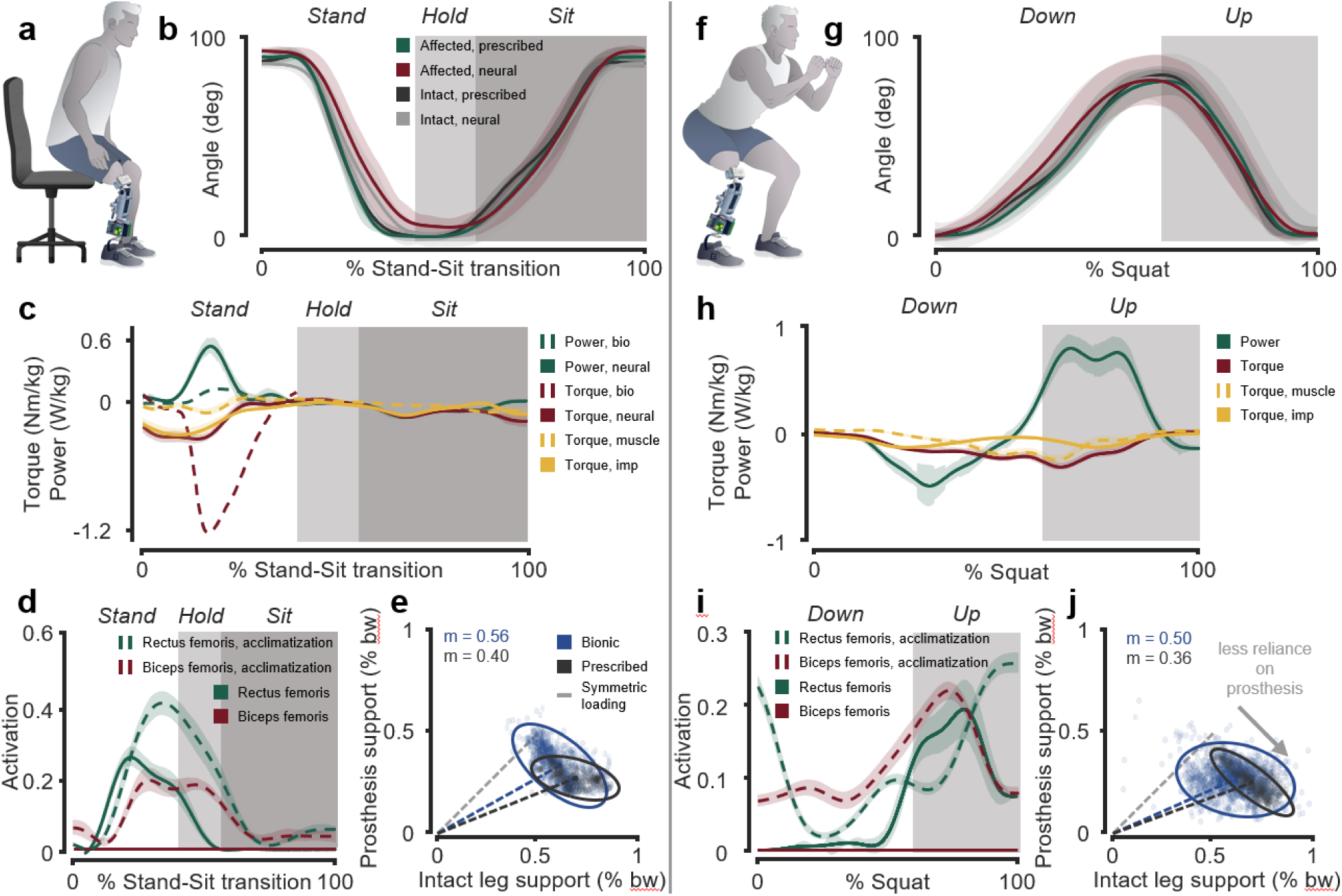
Neural control of sit-to-stand and squatting tasks. Experimental setup for **a** sit-to-stand and **f** squatting tasks. Knee angle profiles during **b** sit-to-stand and **g** squatting. Knee joint torque and power profiles during **c** sit-to-stand and **h** squatting, where muscle and impedance torque traces are the components that make up the total knee torque. Bio represents mean values from literature for healthy adults. Imp represents impedance (baseline) control torque contribution, and muscle represents the contribution from the neural control. Rectus femoris and biceps femoris activation across repetitions during **d** sit-to-stand and **i** squatting. Dashed lines represent the initial 5-minute acclimatization period with the device, and solid lines represent the formal data collection. Shaded regions indicate different task phases. Error bands represent the standard error across repetitions. Load distribution between the prosthetic and intact limbs during **e** sit-to-stand and **j** squatting. Each point represents a time sample. The dashed grey line indicates perfect interlimb symmetry (slope = 1), dashed colored lines show the mean support slope for each device, and solid-colored contours denote the 95% confidence ellipses. The neurally-controlled leg increased prosthesis-side support during both sit-to-stand and squatting compared to the prescribed device.

### Squatting

During repeated squat trials (Fig. 3f), the participant was able to achieve coordinated, full-range knee flexion and extension using both devices. Joint trajectories remained smooth and bilaterally symmetrical (Fig. 3g). The neurally-controlled prosthesis provided a peak power of 0.8 W/kg (Fig. 3h). Throughout initial practice with the squatting task, muscle coordination evolved from dual flexor-extensor activation to exclusive use of the rectus femoris, mirroring control refinement observed during sit-to-stand transitions (Fig. 3i). GRF symmetry increased with the neurally-controlled leg compared to the prescribed device (Fig. 3j).

### Level-Ground Walking

The participant demonstrated symmetric gait with the neural prosthesis during treadmill walking at matched speeds (Fig. 4a) [see Additional file 2]. We define the intact limb as the participant’s non-amputated leg and the affected limb as the leg with the transfemoral amputation. On average, muscle activation patterns were dominated by biceps femoris (Fig. 4c). Kinematic analysis showed improved interlimb symmetry with the neurally-controlled leg (Fig. 4d), and torque and power profiles revealed active extension and positive mechanical work during push-off (Fig. 4e-f). Specifically, the peak power with the neurally-controlled leg was 0.13 W/kg, lower than the peak power of 0.24 W/kg in healthy adults [44]. Temporal gait symmetry, calculated using a modified symmetry index, improved with the neural knee (0.93 vs. 0.59 for the prescribed device) (Fig. 4b). In both treadmill and level-ground walking, the preferred walking speeds were similar across both devices (Fig. 4g-h).

**Figure 4:**
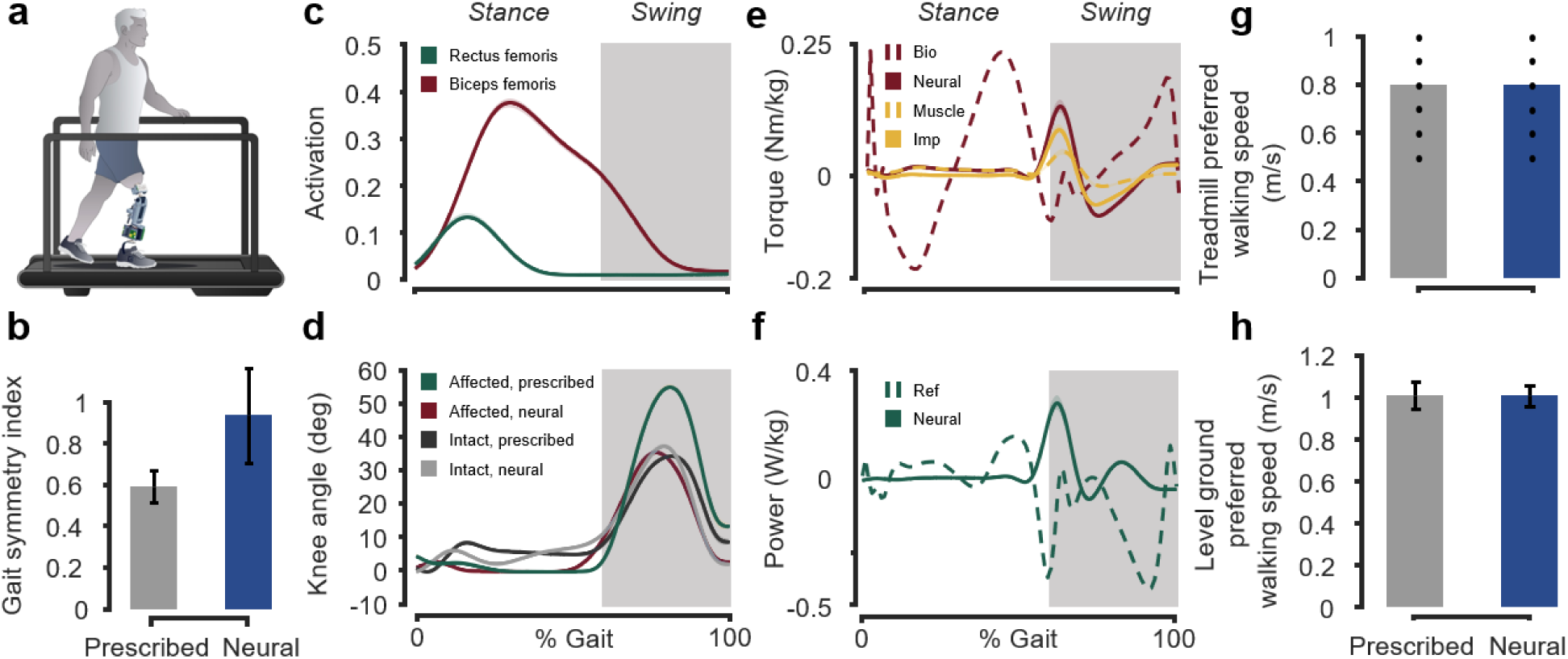
The neurally-controlled leg improves gait symmetry during treadmill walking. **a** Illustration of the experimental setup for treadmill walking with the neural prosthesis. **b** The temporal gait symmetry index improved under neural control compared to the microprocessor knee. **c-f** Knee muscle activation, joint angles, desired torque, and desired power profiles. Bio represents mean values from literature for healthy adults. Muscle and impedance torque traces are the components that describe the total knee torque. Shaded regions in c-f indicate the swing phase. Error bars represent standard error across trials. **g** Treadmill and **h** overground walking speeds were similar across control conditions. Dots represented treadmill speeds evaluated to determine the preferred speed, and error band is the standard deviation.

### Stair Ascent

The participant exhibited improved performance during stair ascent with the neurally-controlled prosthesis compared to their microprocessor knee (Fig. 5a) [see Additional file 3]. Despite similar ascent times across devices (Fig. 5b), only the neural condition enabled a step-over-step pattern, as opposed to the step-by-step pattern used with the prescribed knee. This shift toward more normative gait mechanics was reflected in smooth and continuous knee trajectories of the intact limb, reduced motion variability, and improved joint coordination (Fig. 5d). Muscle activation was timely and consistent with ascending demands both inside and outside the laboratory (Fig. 5c). The peak knee power with the neural leg was 0.17 W/kg both inside and outside the laboratory (Fig. 5e-f), and a value lower than that of young, healthy adults (0.26 W/kg) [45]. These findings demonstrate that the neurally-controlled knee not only restored stepping mechanics but also supported efficient and functional stair navigation. Notably, with both devices, the participant had to use the handlebars for support. Video recordings of outside the laboratory stair ascent further confirm consistent step-over-step behavior in real-world settings [see Additional file 4].

**Figure 5:**
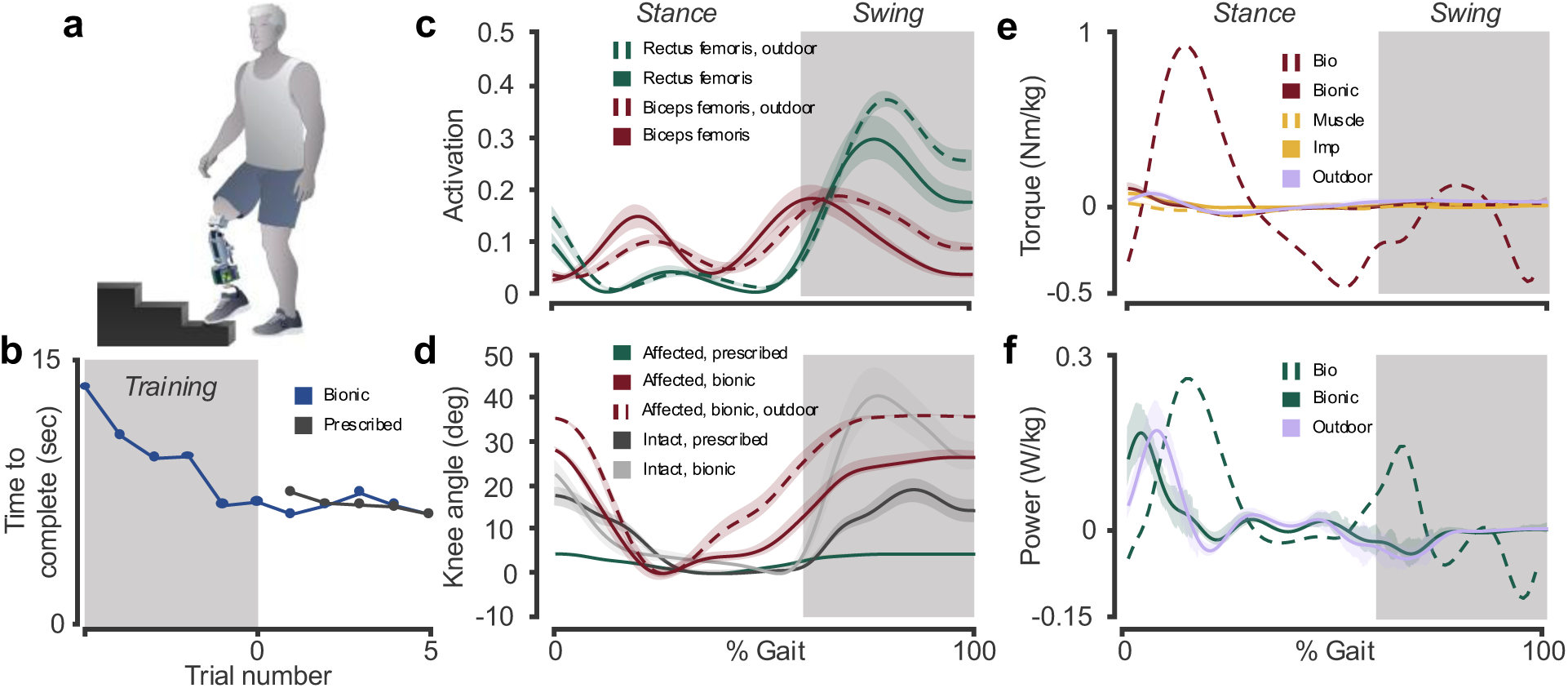
Stair ascent performance and knee joint mechanics. **a** Illustration of the stair ascent task. **b** Trial times show decreasing time to ascend four steps. The shaded region plots the initial 5-minute acclimatization period. **c-f** Knee muscle activation, joint angles, desired torque, and desired power profiles. Muscle and impedance torque traces are the components that describe the total knee torque trace. Bio represents mean values from literature for healthy adults. Shaded regions represent the swing phase of gait, and shaded error bands denote standard error across strides.

### Uninstructed Standing Behavior

During unprompted standing between trials, the participant bore more weight on the prosthetic limb when using the neurally controlled knee. The slope of the regression between prosthetic and intact limb GRFs was higher with the neural leg (0.91 vs. 0.77), indicating greater postural trust outside of explicit task performance (Fig. 6).

**Figure 6:**
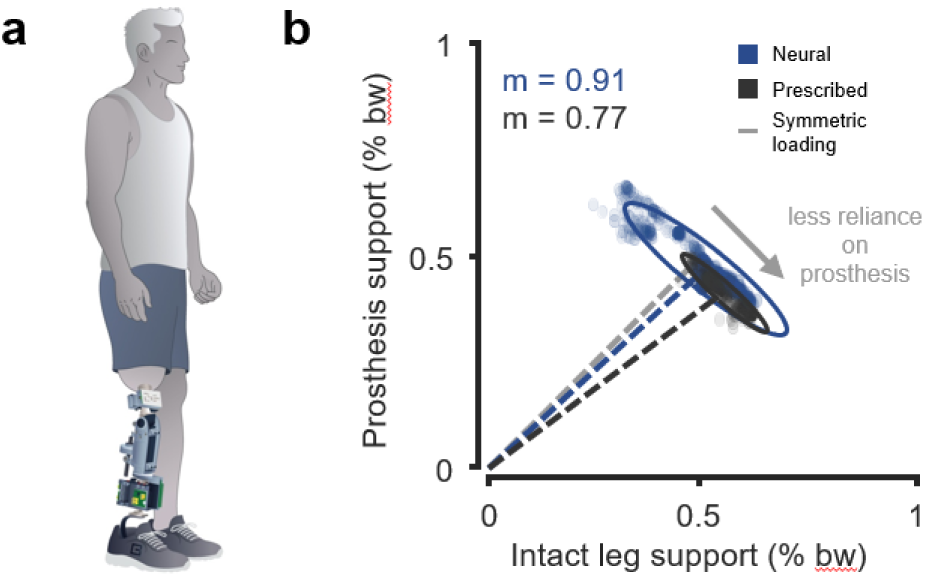
The neurally controlled leg enhances weight-bearing during uninstructed standing. **a** Participant on an instrumented treadmill while awaiting the next task. **b** Prosthesis versus intact leg support (% body weight) during standing intervals. The dashed grey line indicates perfect interlimb symmetry (slope = 1). Dashed colored lines show the mean support slope for each device, and solid-colored contours denote the 95% confidence ellipses. Segment-level support slopes were closer to symmetric loading with the neural leg than with the prescribed device.

### Embodiment

Self-reported embodiment ratings were higher for the neurally-controlled prosthesis across all five measured domains: agency, ownership, body representation, phantom sensations, and proprioception (Fig. 7). Large improvements were noted in ownership and body representation, indicating a greater sense of integration of the prosthesis into the body schema. Subjective comments reinforced these trends, with the participant noting that the neurally-controlled leg “felt like my leg” and “helped [him] more.” Higher phantom sensation scores reflected greater non-painful awareness and integration of the missing limb, rather than increased phantom pain.

**Figure 7:**
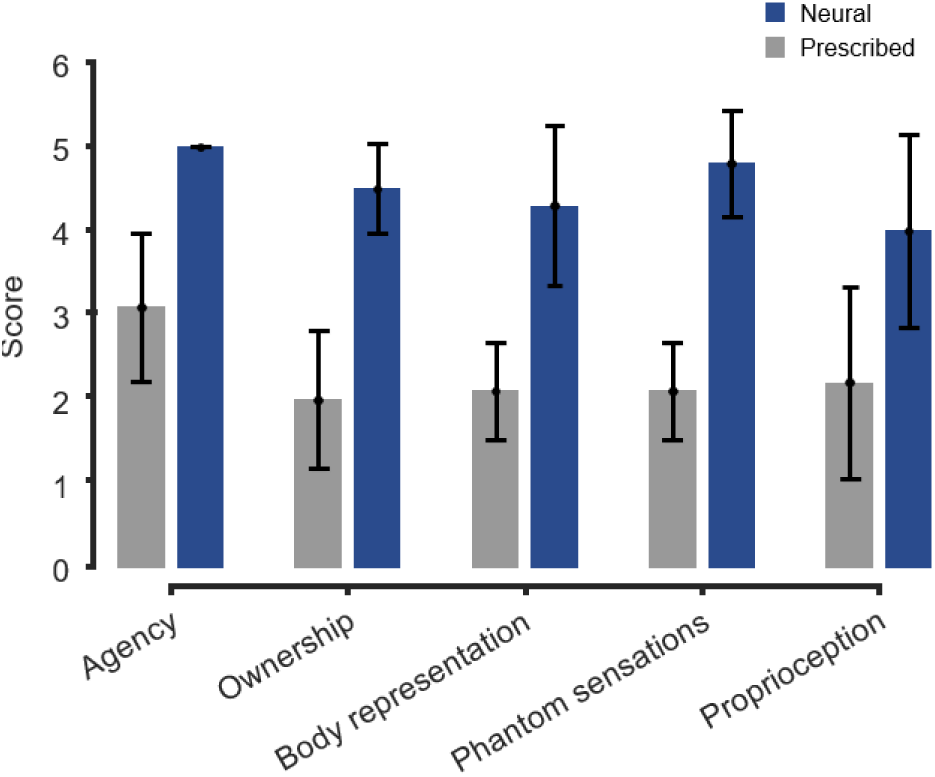
Self-reported embodiment scores were higher for the neurally-controlled prosthesis across all domains. Bar plot shows mean ± standard deviation scores for five embodiment constructs: agency, ownership, body representation, phantom sensations, and proprioception. Scores were rated on a 5-point Likert-type scale (1 = hardly ever, 5 = almost always).

## Discussion

This study demonstrates that a neurally-controlled prosthesis has the potential to significantly enhance functional performance over microprocessor-controlled devices. It explores how bone-anchored, neurally-controlled systems may improve mobility and the sense of embodiment for people with transfemoral amputation. At the transtibial level, prior work has shown that individuals with AMI reconstruction exhibit improved prosthesis control and more biomimetic gait when using neurally-controlled devices compared to individuals with conventional amputations [34], [35]. More recently, a study demonstrated that people with transfemoral amputation with AMI and osseointegration had superior obstacle avoidance and greater prosthesis embodiment than people with a standard amputation [36]. In comparison, powered knees without direct neuro-modulation have also been shown to increase weight bearing symmetry, gait symmetry, and obstacle avoidance [40], [46], [47]. However, these studies did not evaluate the embodiment of the powered and micro-processor legs. In this case study, powered actuation and neural control are intrinsically combined, and our goal was to evaluate the effects of combining a bone-anchored hardware platform with continuous neural control. The presented system appears to afford a higher ceiling for functional performance and body–prosthesis integration than the user’s prescribed microprocessor prosthesis. Future studies should evaluate the contributions from the powered actuation and neural control on these increases. Given the large number of individuals living with conventional amputations, it is clinically important to determine whether more surgically invasive procedures and neurally-controlled devices can provide the significant rehabilitative benefits to justify their continued support and eventual clinical adoption.

In the free-space task, the participant achieved accurate leg positioning using proprioceptive input without any practice with the prosthesis, demonstrating that continuous neural control can enable effective joint modulation without reliance on visual feedback. In both conditions, errors at low and mid targets were larger than the high target, likely stemming from mechanical friction within the actuator which may have limited responsiveness during small, precise movements. Although noise-cancelling headphones were used to attenuate auditory cues, residual actuator noise may still have been perceived and could have contributed to performance or learning effects [48]. Still, the ability to volitionally modulate joint angle under blindfolded conditions supports the efficacy of the neural interface in enabling graded, intuitive control.

During functional tasks, the neurally-controlled leg restored more symmetric kinematics and improved temporal coordination between limbs. Knee flexion patterns during level-ground walking were closely matched between limbs, and the temporal symmetry index increased under neural control. However, stride-to-stride variability in level-ground walking remained higher with the neurally-controlled prosthesis than with the prescribed device, suggesting that gait patterns were still stabilizing during the relatively limited exposure to the novel system. As a result, these temporal symmetry findings demonstrated preliminary evidence that neural control can shift the average pattern toward symmetry, rather than proof of fully stabilized gait. Stair ascent illustrated a similar pattern. The participant consistently used a step-over-step strategy with the neural knee, while the prescribed device required a step-by-step pattern, but the overall stair ascent time was not significantly lower with the neural leg. With additional training, the observed downward trend of stair ascent time may continue, netting improved stair ascent time with the neural device. In outdoor environments, the participant was able to walk smoothly with the neurally-controlled knee. The neural control strategy maintained stable kinematics and joint coordination without the need for manual mode switching, supporting its real-world applicability. Together, these results indicate that neurally-controlled prostheses have the potential to restore more balanced and natural joint trajectories across both steady-state and high-demand tasks, while highlighting the need for longer-term studies of gait variability. These findings are consistent with prior reports that powered knee and knee-ankle prostheses can improve gait symmetry, weight-bearing, and stair traversal in comparison to microprocessor devices [40], [46], [47].

The neural knee facilitated volitional generation of positive power during sit-to-stand, squatting, and stair ascent. In these tasks, peak power was within the range observed in healthy adults, suggesting that the device can restore biomimetic behaviors, despite the user being significantly older than the adults used to generate biological datasets. However, during walking, positive and negative power profiles deviated from able-bodied datasets, likely reflecting compensatory mechanics and the absence of a powered ankle. For example, the positive power right before the transition into swing is the knee compensating for a lack of an ankle that would typically provide push off. In healthy gait, the knee typically performs negative work during this phase; however, in the current system, the knee was required to generate positive work to offset the absence of ankle push-off. These findings highlight the capability of the system to deliver sufficient energy for ambulation, but that powered ankle actuation may be critical for restoring full biomimetic gait.

When standing without prompted, the participant distributed weight more evenly between limbs with the neurally-controlled knee, suggesting natural, unconscious engagement of the prosthesis even outside explicit task demands. These uninstructed intervals were drawn from short standing periods immediately preceding walking and squatting trials, and thus reflect short, sampled postures rather than a continuous, standardized standing task. The observed cluster of intervals with greater prosthetic-side loading in the neurally-controlled condition may therefore reflect specific episodes in which the participant chose to rest more heavily on the prosthesis before tasks, rather than a fully generalizable standing strategy. Nevertheless, the tendency toward more balanced loading in these intervals is consistent with the broader pattern of improved symmetry and suggests that the participant may be willing to rely on the neural prosthesis in low-demand scenarios.

The participant adapted rapidly to selective muscle activation. For example, during sit-to-stand and squatting exercises, the participant chose to back-drive the prosthesis into knee flexion rather than actively contracting the biceps femoris. This strategy reflects increased trust in the device’s passive backdrivability and mechanical stability. By relaxing the biceps femoris during the ascent phases, the participant was able to more precisely modulate extension torque because of the reduced co-contraction. This pattern of task-specific muscle activation suggests that the participant learned to exploit the mechanical characteristics of the prosthesis in a way similar to able-bodied neuromuscular coordination, where antagonistic muscles are selectively inhibited to permit efficient joint extension.

Self-reported embodiment scores were higher across all assessed domains when the participant used the neurally-controlled prosthesis. Pronounced gains were observed in ownership and body representation, indicating that the neural prosthesis was perceived as more integrated into the participant’s body schema; scores in these areas are higher than in prior studies [36], which may be a secondary effect of more mature hardware. Higher scores for phantom sensations and proprioception further suggest that the neurally-controlled leg may enhance sensory awareness and engagement with the residual limb. Despite some variability in item-level responses, the consistent pattern across all five domains underscores the potential for neural control to meaningfully enhance prosthetic embodiment.

The system did not require recalibration or parameter tuning once initially configured. The participant completed all functional tasks using a single control configuration across multiple sessions and environments, including outdoor settings. This consistency may be a result of the high signal-to-noise ratio and stability of the implanted electrodes, which remained unaffected by sweat, movement, or limb displacement. In contrast, surface EMG systems often necessitate recalibration and are susceptible to signal degradation over time [49]. The signal fidelity and reliability of implanted electrodes represent a significant practical advantage for the real-world deployment of neural prosthesis control systems.

While this study demonstrated the feasibility and promise of myoneural control for powered knee prostheses, several limitations must be acknowledged. First, this was a single-participant case study, which limits generalizability and precludes broader statistical inference. Additionally, the participant had prior experience with neurally-controlled prosthetic systems, which likely facilitated adaptation and may not reflect the learning curve encountered by first-time users. The uninstructed standing analysis was based on brief standing intervals that occurred before specific functional tasks, which could be subject to sampling bias. As such, the observed episodes of increased prosthetic loading in the neurally-controlled condition may not reflect a general standing strategy. Future studies should include a larger and more diverse cohort, employ validated embodiment instruments where possible, and consider third-party administration of questionnaires to further minimize response bias.

The evaluation of certain high-demand tasks, such as stair descent, was constrained by hardware limitations. Specifically, the eOPRA implant imposed a ±70 Nm torque limit, which restricted the prosthesis’s ability to deliver sufficient damping torque for safe stair descent. Even during stair ascent, the participant required the use of handrails to compensate for limited torque assistance. These findings suggest that torque constraints imposed by the osseointegrated implant may restrict performance in high-load activities. Future studies should include participants without OPRA-based torque limitations to more fully evaluate stair navigation.

This study investigated the capacity of a neurally-controlled leg with combined intrinsic and neural control to support ambulation and high-demand tasks in unstructured, real-world environments. Recent work has introduced a unified knee–ankle controller capable of adapting to varied terrain during outdoor ambulation [50]. While this system demonstrated robust environmental adaptability, it lacked volitional control and may not support the same degree of embodiment as a neuro-modulated device. In contrast, the neurally-controlled leg presented in this study enabled continuous, user-driven modulation of joint torque based on efferent muscle signals. Integrating neural control with unified, terrain-adaptive frameworks may represent a promising direction for future prosthetic systems, offering both environmental robustness and intuitive, embodied user interaction.

## Conclusions

This study provides evidence that a neurally-controlled prosthesis can restore volitional control, functional capability, and embodied use of a prosthetic limb in individuals with transfemoral amputation. Compared to the participant’s microprocessor knee, the neural prosthesis improved biomechanical symmetry, load distribution, and task efficiency, while also enabling greater user confidence and weight-bearing. Notably, the neural leg enhanced subjective embodiment, as reflected by increased self-reported scores in domains of agency, ownership, and body representation. These findings suggest that neurally-controlled prostheses have the potential to achieve not only enhanced functional performance but also deeper integration into the user’s body schema.

## Data Availability

All data are available in the main text or the supplementary materials.

## List of abbreviations

AMI: Agonist-antagonist myoneural interface
eOPRA: electronic Osseointegrated Prostheses for the Rehabilitation of Amputees
ROM: Range of motion
ADC: Analog-to-digital converter
IMU: Inertial measurement unit
CAN: Controller area network
CANOpen: CAN object exchange protocol
LiPo: Lithium-polymer (battery)
DC-DC: Direct-current to direct-current
DOF: Degree-of-freedom
EMG: Electromyography
FSM: Finite state machine
GRF: Ground reaction force
PROM: Patient-reported outcome measures.

## Supplementary Material

**Additional file 1.** Supplementary Materials: Methods, Figures, and Tables.

**Additional file 2.** Movie of treadmill walking with both devices.

**Additional file 3.** Movie of stair ascent of 4 steps with both devices.

**Additional file 4.** Movie of stair ascent inside the Media Lab with the neurally-controlled prosthesis.

## Acknowledgments

We gratefully acknowledge our clinical collaborators K. Clites, C. Sullivan, and G. Carson for their invaluable support. The authors would also like to thank M. Nawrot and D. Lee for their work building and maintaining the motion capture system. We also thank N. Goderdzishvili for their artistic contributions.

## Authors’ contributions

JM designed the experiments, conducted data collection and analysis, prepared figures, and wrote the main manuscript text. HH supervised the study. All authors read and approved the final manuscript.

## Availability of data and materials

All data are available in the main text or the supplementary materials.

## Ethics approval and consent to participate

Experiments were conducted with informed consent at the Massachusetts Institute of Technology (MIT) Media Laboratory under COUHES protocol #2503001589.

## Consent for publication

All subjects gave their permission through written informed consent to publish individual data as picture or video.

