## Supplemental Methods for "Design and Evaluation of a Bone-anchored, Neurally-controlled Knee Prosthesis"

John A. McCullough *et al.*

The PDF file includes:

Supplementary Methods

Figs. S1 to S7

Tables S1 to S2

Other Supplementary Material for this manuscript includes the following:

Movies S1 to S3

### Supplementary Methods

#### Embedded Control Architecture

The control system of the prosthesis centers around a Toradex Verdin iMX8M Plus computer (Fig. S1), which runs a multithreaded control stack on embedded Linux. The Verdin interfaces with the motor driver and peripheral sensors via CAN, universal serial bus (USB), and analog channels to support real-time control and data acquisition. Motor position is tracked using a high-resolution incremental encoder, and joint angle is measured via an absolute encoder, both connected to the Ingenia Everest XCR motor driver. The driver communicates with the Verdin computer at 1 kHz over CANOpen. Sensor inputs include a 6-DOF load cell, an axial load cell, and a custom EMG board. The Verdin polls all sensors at 1 kHz and processes EMG signals in real time to compute muscle activation. These signals modulate torque commands and impedance parameters. This architecture supports synchronized control and data logging across all subsystems, ensuring consistent timing between neural inputs, actuator commands, and sensor feedback.

#### Software Stack

The control software for the powered knee prosthesis is built on a modular, object-oriented architecture running (Fig. S2). The software stack, based on the Talaria framework, operates within a Yocto Project-based embedded Linux environment and supports deterministic, multithreaded execution at 1 kHz. Sensor polling, control computation, and motor command updates run concurrently in soft real-time, ensuring consistent low-latency performance. All sensors are interfaced through dedicated software modules and polled at 1 kHz. The motor driver is controlled via CANOpen using the open-source Lely library. Desired joint torques are computed from neural inputs and mapped to motor current commands, which are transmitted to the driver each cycle. The CANOpen network is configured using autogenerated device files, enabling quick reconfiguration and minimizing bus overhead.

At runtime, the system launches multiple threads: sensor polling and control logic, EMG acquisition, data logging, and external communication. In the control logic, an FSM selects the active controller based on gait phase, inferred from peripheral sensors, and then computes the desired torque based on impedance parameters. Logging is handled asynchronously and records time-stamped data at 1 kHz for offline analysis.

Safety is ensured through automated shutdown protocols that send stop commands, reset control variables, and close communication channels on termination. This architecture enables stable high-frequency control while maintaining flexibility for expansion to multi-joint or multi-user deployments.

#### Finite-State Machine

An FSM governs the high-level control transitions of the knee prosthesis, differentiating between stance and swing phases of gait. Transitions between states are determined using ground contact information derived from a 6-DOF load cell mounted distal to the prosthesis (Fig. S3). When the load cell detects unloading of the limb, the controller transitions from stance to swing; conversely, limb loading triggers a transition from swing to stance.

#### Intrinsic Controller

The intrinsic controller provides baseline knee behavior using impedance control modulated by an FSM that differentiates between stance and swing. The FSM does not perform explicit classification of specific tasks, such as level-ground walking, stair ascent, or descent. Instead, it provides task-general flexion and extension dynamics that can adapt across various functional activities.

**Neural Controller**

The powered knee prosthesis control incorporates a neural layer designed to enable the user to volitionally control the knee, published previously [36]. EMG signals are acquired from flexor and extensor muscles in the residuum and processed to compute a desired knee torque in real time. This neural control layer operates in parallel with the intrinsic FSM controller and contributes continuously to the joint torque without reliance on terrain or gait classification (Fig. S4).

Muscle activation signals are mapped to adjust joint stiffness, damping, and equilibrium angle. Specifically, logistic functions are used to compute the angular setpoint as a function of the difference between flexor and extensor activations, with additional modulation based on recent activation history to provide smooth transitions. Stiffness and damping values are determined using distributive and affine mappings of muscle activations, allowing for physiologically relevant adjustments in joint impedance. The result is a user-driven modulation of knee behavior, enabling the wearer to augment, fine-tune, or override baseline knee dynamics in response to task demands or environmental conditions.

**Torque Control Loop**

The desired torque is passed to a low-level torque control loop responsible for accurately regulating knee torque (Fig. S5). A closed-loop torque control system is implemented using a combination of feedforward and feedback pathways [36]. The closed-loop controller achieves a torque bandwidth of approximately 10 Hz, enabling accurate tracking of commanded joint torques. Given that the dominant frequency content of human locomotion is typically below 6 Hz, a 10 Hz closed-loop what is bandwidth is sufficient to stably reproduce the relevant joint dynamics [51]. Feedforward terms compensate for known system dynamics, while feedback terms correct for unmodeled disturbances and improve torque tracking accuracy through a proportional-integral controller.

**Joint Encoder**

The original joint encoder (AMS AS5048B, $I^2C$ protocol, 14-bit resolution) was replaced due to electromagnetic interference (EMI) vulnerability, which caused intermittent communication failures. The updated magnetic absolute encoder (RM08, RLS, Ljubljana, Slovenia) uses a synchronous serial interface (SSI) with differential signaling, providing substantially greater EMI immunity. Although the RM08 offers a lower nominal resolution (12-bit, 0.088°/count), this is more than sufficient for knee joint sensing in this application. At the closed-loop bandwidth of the knee of 10 Hz and a 1 kHz control loop, the in-band position quantization noise is approximately 0.003° RMS, over an order of magnitude below typical human joint position variability (0.1° RMS) and well beneath the error threshold that would affect velocity estimation or torque control. Post-integration, no packet loss or communication errors were observed, and control performance was noticeably smoother during high-speed, high-torque tasks.

**Motor Encoder**

The motor position is measured using a RLS RLM2 magnetic rotary encoder from the previous iteration of the knee [37]. The encoder achieves an effective resolution of 57,600 counts per revolution. This incremental encoder interfaces directly with the Ingenia Everest XCR motor driver via RS-422 differential inputs. The encoder enables real-time computation of rotor angle and velocity within the servo drive, supporting stable and accurate torque regulation during dynamic movements.

**Axial Load Cell**

A compact axial load cell (LCM300, Futek) was integrated into the actuator assembly to measure actuator-generated forces. This sensor, retained from the original system [79], has a maximum capacity of 4450 N (1000 lbf) and exhibits high linearity and low hysteresis. It is connected to a 32-bit onboard ADC, allowing high-resolution force measurements suitable for real-time feedback control.

**6-DOF Load Cell**

To improve ground contact detection and reduce latency, the previous force-sensitive resistor (FSR) array was replaced with a 6-DOF load cell (M3564F1, Sunrise Instruments). The load cell provides measurements of ground reaction forces (Fx, Fy: 2500 N; Fz: 5000 N) and moments (Mx, My: 200 Nm; Mz: 100 Nm) with minimal crosstalk and hysteresis. It communicates with the onboard computer via RS232 over USB at 600 Hz. Mounted in-line between the actuator and the passive ankle, the load cell directly captures joint torque and axial force. Ground contact detection latency was reduced from 25–30 ms (FSR-based) to 3–5 ms, improving phase transition detection. The real-time availability of torque measurements enables implementation of torque-feedback control strategies.

**Power Distribution Board**

A custom-designed power management board was developed to reliably deliver and regulate electrical power for both high-load actuation and low-voltage control electronics (Fig. S6). The system is supplied by two 6S1P 22.2 V, 1000 mAh lithium polymer (LiPo) batteries connected in series, providing a nominal 48 V power rail for the actuator.

To supply low-voltage electronics, a 12 V isolated DC-DC converter (Murata UEI15-033-Q48N-C) steps down the 48 V input, delivering regulated power to components including the onboard computer and load cell data acquisition system. A high-side mechanical switch is integrated into the 12 V line, enabling manual disconnection of the low-voltage rail for safety during maintenance or emergency situations. An additional emergency stop on the board triggers the motor driver's safe torque off mode.

High-voltage delivery to the motor driver is controlled by a solid-state relay (Sensata-Crydom CMX60D20), actuated via a general-purpose input/output signal from the Verdin onboard computer. This setup allows the motor driver to be selectively powered down by software control, enabling safe startup and shutdown routines and conserving energy during idle periods. The relay is rated for 20 A, so software limits are implemented to limit the maximum allowable current to ±20 A to prevent damage to the relay. In practice, typical current demands during locomotion remain well below this threshold.

A bulk capacitor is installed on the 48 V rail to buffer high transient loads from the actuator, minimizing voltage sag during rapid current draws and maintaining system stability. PCB trace widths were designed based on measured root mean square currents from field experiments, with thermal limits set to ensure temperature rise remains below 50°C under typical locomotion conditions.

The battery voltage is continuously monitored using an onboard voltmeter (Adafruit 705), powered by a dedicated 12 V buck converter (Recom Power R-78HB12-0.5/W). This allows users to track battery levels in real time and replace batteries before they reach deep discharge. Visual feedback is provided by three onboard LEDs, indicating the status of the 12 V rail, 48 V battery supply, and motor driver power line.

Additional features on the second layer of the board expand sensing and communication capabilities. A compact IMU (Bosch BMI270) is integrated to support onboard kinematic state estimation. A high-resolution 32-bit ADC (TI ADS1262IPW) is included to provide analog input capability in cases where the carrier board lacks high-resolution ADCs. Finally, two CAN transceivers (Microchip MCP2562) are incorporated to enable CAN communication with carrier boards that do not include integrated transceivers, such as the Mallow carrier board (Toradex AG, Horw, Switzerland).

Thermal imaging data collected using a FLIR camera (Teledyne Technologies, Thousand Oaks, CA) during continuous operation validated that the board did not exceed 50 °C. These measurements validate the board's thermal design and current handling capacity. Overall, this power management architecture ensures stable operation of both high-power and low-voltage subsystems, while offering enhanced safety, monitoring, and control features critical for experimental use and field deployment.

**Motor Driver Heat Sink Thermal Analysis**

A modular heat sink was designed to provide reliable thermal dissipation under high-load conditions typical of dynamic locomotion tasks. The heat sink was fabricated from Aluminum 6061-T6 (Aether Manufacturing Inc., Brooklyn, NY). A figure illustrating the thermal design process and heat transfer calculations is provided in Figure S7.

**Embodiment Questionnaire**

At the beginning and end of the testing week, the participant completed a questionnaire reflecting on their experiences with their prescribed and neural devices (Table S2). The set of questions administered in this questionnaire was designed by the researchers and not validated externally. The questionnaire consisted of close-ended questions grouped into three embodiment domains: agency, ownership, and body representation. It also included questions on phantom sensations and proprioception. Each question used a 5-point Likert scale, ranging from "hardly ever" to "almost always." Responses were numerically coded from 1 to 5.

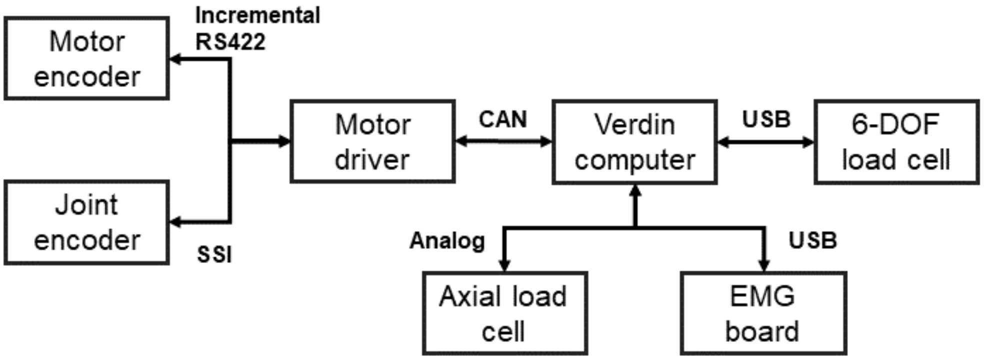


Figure S1: System architecture illustrating core communication pathways in the prosthesis control system. The motor encoder and joint encoder transmit position to the motor driver. The Verdin computer communicates with the motor driver while simultaneously interfacing with peripheral sensors. This architecture enables synchronized real-time data acquisition and motor control.

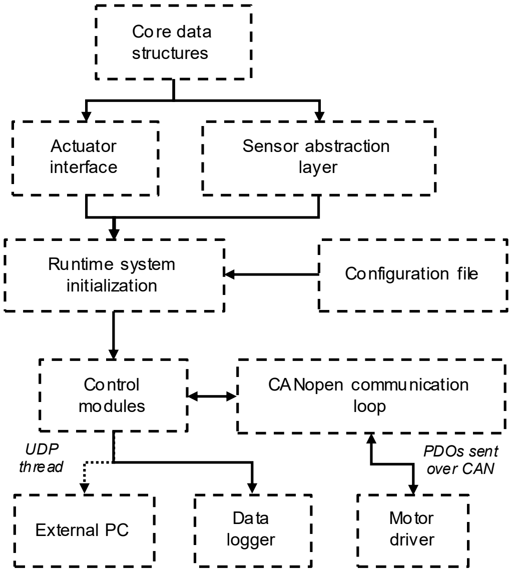


Figure S2: Object-oriented software architecture of the powered knee prosthesis. The architecture is organized into modular layers.

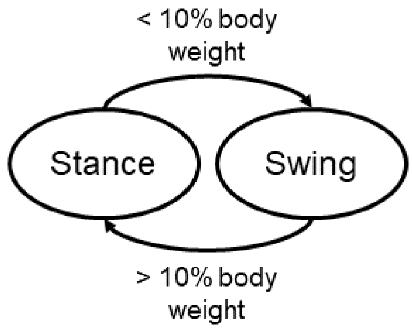


Figure S3: Finite state machine for gait phase detection. The controller switches between stance and swing phases based on vertical ground reaction force thresholds. A transition from stance to swing occurs when load on the prosthesis drops below 10% of body weight, and swing-to-stance transitions occur when load exceeds 10% of body weight.

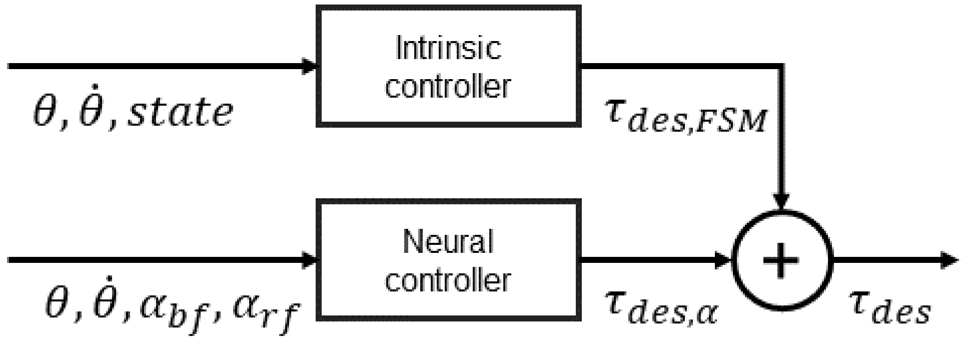


Figure S4: Block diagram of the parallel control architecture. The desired joint torque is calculated as the sum of two parallel control pathways. The intrinsic controller generates task-specific torque based on joint angle, joint velocity, and gait state. The neural controller produces volitional torque commands based on joint kinematics and processed EMG signals from the muscles. This combined architecture enables both automatic task execution and user-driven modulation of the prosthetic knee.

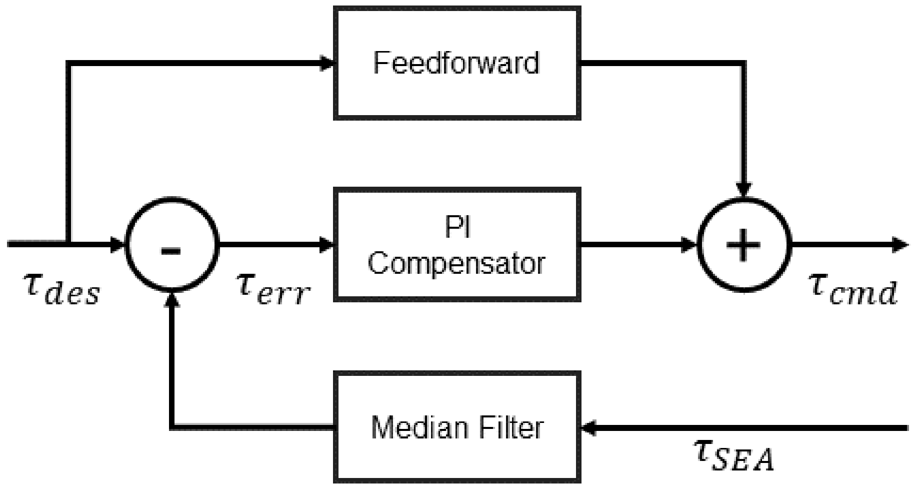


Figure S5: Closed-loop torque control architecture for the prosthetic knee. The desired torque is compared to the measured torque to compute the error signal. This error is processed through a proportional-integral compensator, while the measured torque is first passed through a median filter to reduce noise. A feedforward pathway adds additional responsiveness based on known dynamics. The resulting output command torque ensures accurate and stable joint actuation.

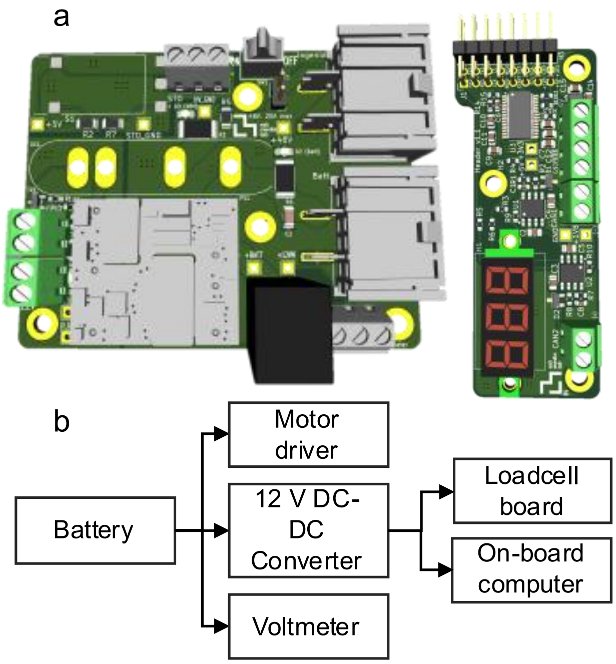


Figure S6: Custom power management system for the powered prosthesis. **a** Renderings of the custom-designed power distribution board. **b** Block diagram illustrating power flow through different components. This system provides centralized, regulated power distribution to all critical components.

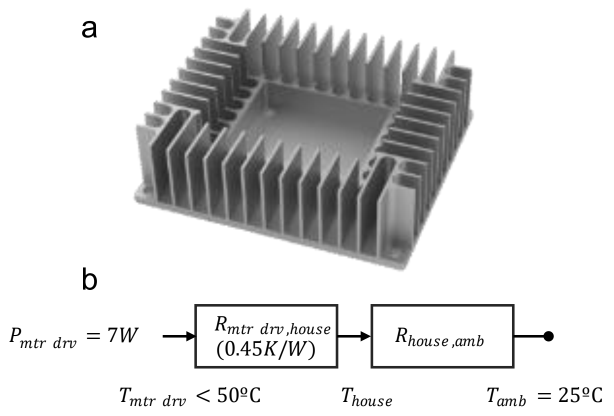


Figure S7: Heat dissipation strategy for the motor driver. **a** Three-dimensional rendering of the heat sink enclosure. **b** Thermal resistance model used to estimate motor driver temperature under a 7 W load, showing conduction to the housing and convection to ambient air.

Table S1: Mass distribution of the knee prosthesis

| **Item** | **Mass (g)** |
|---|---|
| Structural components | 650 |
| Motor | 550 |
| Spring | 180 |
| Ball-screw and in-line loadcell | 150 |
| Electronics | 440 |
| Fasteners | 80 |
| Batteries | 350 |
| Case | 200 |
| Total | 2600 |

Table S2: Embodiment Questionnaire

| Theme | Question |
|---|---|
| Agency | My leg moves the way I want my leg to move |
| | My leg only moves when I want my leg to move |
| | If my leg moves, it feels like I am moving it |
| | I am in control of the movement of my leg |

| | |
|---|---|
| | When I want to perform a movement, I trust my leg to perform my intended movement |
| | I feel like I initiate the movements of my prosthetic leg |
| | I can make my prosthetic leg move exactly when I want it to |
| | I do not feel surprised by the movements of my leg |
| | The movements of my leg feel intentional |
| | When my leg moves, it matches my intended action |
| Ownership | I am able to sense where my leg is in space |
| | I am able to sense how the joints of my leg are positioned |
| | With my eyes closed, my leg feels like my non-amputated limb |
| | When I move it and then open my eyes, my leg is located where I expected it to be |
| | The posture of my leg corresponds to that of a non-amputated leg |
| | I can tell when my leg has moved, even without looking |
| | I have a constant awareness of the presence of my leg |
| | My leg feels like it is the correct size and length for my body |
| | I do not have to look down to locate my leg |
| | My leg feels like a natural part of my body |
| Body Representation | My leg feels like it belongs to me |
| | My leg feels like it is a part of me |
| | When I look at my leg, I feel like I am looking at a part of myself |
| | I feel as if the leg is my leg |
| | My leg feels physically and naturally integrated with my body |
| | I feel attached to the leg |
| | My leg feels foreign, like it does not belong to me |
| | My leg feels like a tool or device rather than part of my body |
| | I am relieved when I remove the leg |
| | I feel uncomfortable when I am not wearing the leg |
| Proprioception | I can tell where my leg is in space without looking |
| | I can sense how my knee is positioned |
| | I can sense when my leg is moving |
| | I know how far I’ve moved my leg during a motion |

| | |
|---|---|
| | When I move my leg, I can feel the movement from within |
| | Even with my eyes closed, I can picture the orientation of my leg |
| | I can feel whether my knee is bent or straight |
| | I can feel how fast my leg is moving |
| | I am aware of my leg's posture during daily activities |
| | When my leg is moved passively, I can tell which direction it's going |
| Phantom Sensations | I can feel the presence of my missing limb |
| | I am aware of the shape or outline of my phantom limb |
| | I can feel movement in my phantom limb |
| | I can intentionally move parts of my phantom limb |
| | My phantom limb feels like it is positioned naturally |
| | I can feel specific parts of my phantom limb (e.g., toes, foot, knee) |
| | The position of my phantom limb changes throughout the day |
| | I can use my phantom limb to help visualize or guide movement |
| | My phantom limb feels like a part of my body |
| | I can feel sensory experiences (e.g., pressure, temperature, texture) in my phantom limb. |
